# Evaluation of an Integrated Digital and Mobile Intervention for COPD Exacerbation

**DOI:** 10.1101/2025.02.13.25322246

**Authors:** Laurel O’Connor, Biqi Wang, Zehao Ye, Stephanie Behar, Seanan Tarrant, Pamela Stamegna, Caitlin Pretz, Apurv Soni

## Abstract

**Background:** Chronic obstructive pulmonary disease (COPD) is a leading cause of morbidity and healthcare utilization, with frequent exacerbations contributing to emergency department visits and hospitalizations. This study evaluates a multimodal, community-based digital health intervention’s association with changes in acute care utilization among patients with COPD to develop preliminary estimates of intervention effects.

**Methods:** In this decentralized, nonrandomized pilot clinical trial, participants with moderate to severe COPD were offered biometric monitoring, symptom tracking, on-demand MIH services, and a digital pulmonary rehabilitation program. Outcomes were compared between intervention participants and a weighted synthetic control group using full optimal matching. Weighted odds ratios derived from regression models were used to estimate intervention effect size. The primary outcome was hospitalization during the study period. Secondary outcomes included 30 and 90-day readmission rates, emergency department visits, and hospital length of stay.

**Results:** In total, 88 participants from the intervention arm (mean age 67, 50% female) were compared to a weighted synthetic control of 14,492 participants (weighted mean age 66, 48.7% female). We observed that participants in the intervention arm had a trend toward decreased hospitalization with an OR of 0.69 (CI 0.44-1.03, p=0.066). The intervention was also associated with 61% decreased odds of 30-day readmission after an index admission compared to controls (OR: 0.39, 95% CI: 0.16–0.95, p = 0.04). Trends toward reductions in ED visits and hospital length of stay were also observed.

**Conclusions:** A combined digital and mobile health approach to COPD management was associated with reductions in acute care utilization. These findings support further investigation into hybrid care models to enhance COPD self-management and improve patient outcomes. Future research should evaluate scalability, cost-effectiveness, and long-term clinical impact.

## Introduction

Chronic obstructive pulmonary disease (COPD) poses a significant public health challenge internationally, impacting half a billion individuals worldwide.^1, 2^ In the United States, 16 million individuals have been diagnosed with COPD, representing about 10% of adults 40 and older.^3, 4^ Its hallmark symptoms include breathlessness, chronic cough, and fatigue, which diminish patients’ quality of life and impact their ability to work, participate in social activities, and perform daily tasks.^5^ Additionally, frequent periods of worsening symptoms (exacerbations) often result in ED and hospital stays, which further strain patients and their families.^6^ These exacerbations cause 873,000 ED visits and 700,000 hospitalizations annually in the U.S. and cost 70% of the estimated $50 billion spent annually on COPD.^6, 7^ Up to 50% of patients with COPD over the age of 40 have at least one exacerbation a year, and nearly 20% of those hospitalized for an exacerbation require readmission within 30 days.^1, 5, 8^

To improve disease management, reduce healthcare costs, and improve patient quality of life, proactive strategies are essential to minimize the frequency and severity of COPD exacerbations. Timely recognition of early clinical signs of COPD exacerbation, initiation of appropriate pharmacologic therapy, close symptom monitoring, and participation in pulmonary rehabilitation can help prevent severe disease progression and associated complications^5, 8–11^. However, patients often don’t recognize or act on symptoms of COPD exacerbation until symptoms have become severe.^12–14^ Further, they encounter barriers to care access, including financial stressors, transportation difficulties or geographical distance from healthcare facilities, ambulatory capacity limitations, difficulty navigating the healthcare system, and lack of social support.^14, 15^ Finally, only about 4% of eligible patients actually participate in pulmonary rehabilitation despite strong evidence of its efficacy. ^16, 17^ Thus, ensuring the timely and effective delivery of these evidence-based strategies remains a critical challenge in COPD care.

Interventions that mitigate barriers to COPD care access using mobile and digital resources to make it easier for patients to access physiologic monitoring, urgent clinical evaluation, and treatment show promise for facilitating the expedient detection and treatment of COPD exacerbation.^18–20^ Specifically, home monitoring systems using patient-reported changes in symptoms and wearable sensors that transmit biometric data have been shown to accurately predict clinical deterioration.^18^ Both in-person and remote pulmonology rehabilitation and lifestyle coaching have demonstrated effectiveness in improving clinical outcomes in COPD, including quality of life, functional capacity, and patient-reported dyspnea, which in turn decreases acute care needs.^21, 22^ Finally, mobile Integrated Health (MIH) models, healthcare delivery programs that leverage mobile resources, including specially trained paramedic-level clinicians, to provide preventative and acute care for patients at home, have been shown to increase patient satisfaction, care delivery quality, and decrease healthcare costs.^20, 23, 24^ Individually, these interventions provide incremental solutions to current limitations in COPD care. However, if successfully integrated, there may be potential to develop a unified approach to expediting the recognition, evaluation, and treatment of COPD exacerbation. Few interventions have examined the feasibility or impact of a combined model that integrates these valuable resources into an integrated approach to community-based care for COPD. While such an approach may bring together evidence-based strategies for COPD management, the effectiveness of combining approaches into a single complicated but comprehensive intervention is unknown. We previously described the feasibility of implementing a multimodal, community-based intervention in treating acute COPD exacerbations, the “Healthy at Home” model, which combined remote monitoring, telepulmonary rehabilitation, and mobile integrated health.^25, 26^ The intervention achieved its desired recruitment rate and demonstrated a decrease in COPD-related distress in participants. The objective of the present study is to develop preliminary effectiveness estimates for the clinical impact of the Healthy at Home Intervention.^25, 26^

## Methods

### Setting and Participants

This decentralized, nonrandomized pilot clinical trial was conducted through an urban academic tertiary care center, whose emergency departments collectively manage approximately 200,000 acute encounters annually. A detailed description of the study protocol and intervention characteristics is described elsewhere.^25^ Eligible participants were required to receive healthcare within the affiliated hospital system, be 18 years or older, speak English, and have a documented diagnosis of COPD with a moderate to high risk of hospitalization, as determined by an institutional algorithm. Additionally, participants needed to own a smartphone. Exclusion criteria included inability to provide informed consent, lack of English proficiency, absence of internet access at their place of residence, concurrent enrollment in another clinical trial, or prior participation in any Wellinks pulmonary support program.

Eligibility screening was conducted using the hospital system’s electronic health record (EHR) to identify patients with a 25–50% predicted risk of COPD-related hospitalization within six months. Risk stratification was based on acute care utilization (ED visits and hospitalizations) and COPD-related medication changes over the preceding two years. Recruitment targeted patients in the second to fourth quintiles of this risk distribution to ensure a moderate-to-high admission risk cohort. A sample size of 100 participants was selected to assess feasibility, estimate variability, and refine study protocols in preparation for a larger, fully powered trial.

Participants were initially invited to enroll via email, with non-responders receiving text message follow-ups and paper mailers within two weeks of contact. Those identified as potential participants were also approached in person during hospitalizations or pulmonary clinic visits if upcoming in-person encounters were expected during the study period. Additionally, study flyers were placed in clinic waiting areas to allow participants to self-refer to the study. Recruitment invitations occurred in waves of 500–3,000 patients and were adjusted to ensure a balanced representation across risk and sociodemographic groups.

Interested participants underwent eligibility screening, digital consent, and enrollment via the MyDataHelps app, a custom service that provides a platform for patient-facing instruments in clinical trials. Upon enrollment, a welcome kit was shipped to each participant, including a Fitbit Charge 5, study materials, and instructions for requesting Mobile Integrated Health (MIH) visits. Remote support was provided for smartwatch setup and app navigation via phone or video, based on participant preference. Participants engaged in study activities for six months.

Patients who completed consenting procedures and successfully enrolled in the study were considered the program-treated group. Patients who passed the initial eligibility assessment but were never invited were considered part of the program control group. For the program-treated group, enrollment dates ranged from 10/20/2022 to 2/02/2024. The study was approved by the WIRB-Copernicus Group Institutional Review Board. It is registered at Clinicaltrials.gov (NCT06000696).

### Intervention

The Healthy at Home intervention included several complementary elements, including biometric monitoring, symptom tracking, mobile acute care services via an institutional MIH program, and optional digital pulmonary rehabilitation by Wellinks. Patients who opted into the Wellinks Virtual Pulmonary Rehabilitation Program were considered as Wellinks treated group. Supplementary Table 1 summarizes each component and its interaction with other study constituents.

### Clinical outcomes

The primary clinical outcome was hospitalizations during the study period. Secondary outcomes included hospital readmissions within 30 and 90 days, number of ED visits, and annual average hospital length of hospital stay. We also performed a difference-in-differences analysis, evaluating the difference in the number of hospitalizations, ED visits, mean length of stay, and 30 and 90-day readmissions in the pre and post-intervention period with a comparison between the intervention and control group.

Hospital utilization indices were extracted from patient electronic health records (EHR) under the OMOP Common Data Modal. Acute care encounters were tracked in the visit occurrence database with specific concept identifications: inpatient visits were coded by 9201, outpatient visits were by 9202, and emergency department (ED) and inpatient visits were by 262. Readmission days were calculated by the time differences between the discharge dates and the readmit dates among inpatient and ER visits. We also calculated hospital length of stay (LOS), which was the days within the hospital for every inpatient visit.

All hospital utilization indices were censored 1 year after the index date, which was the actual enrollment date for the treated group. The first enrollee’s enrollment date was set as the index date for the controls. The pre-intervention period was defined as less than 1 year before the index date, and the post-intervention period was within 1 year after the index date. Binary outcomes were any readmissions within 30 days of hospital admission, readmissions within 90 days, any inpatient visit during the study period, and any ED visit during the post-intervention period. Continuous outcomes included average LOS after the index date, the difference in the number of 30 and 90 days readmission changes between pre and post-index date, the difference in number of 90 days readmissions, the difference in number of ED visits changes, the difference in inpatient visits changes, and difference of average LOS changes between pre and post-intervention periods (i.e., mean LOS before enrollment date minus the mean LOS after enrollment date).

### Other measurements

Demographic factors, including age at the enrollment, sex assigned at birth (male or female), race (White, Black/African American, Other), and ethnicity (Hispanic or non-Hispanic), were abstracted from the EHR. The unweighted Charlson Comorbidity Index (CCI) score was generated as the number of health conditions (up to 14 conditions) before the enrollment date.

### Statistical analysis

To optimally evaluate the effectiveness of the program intervention in a nonrandomized trial, we used optimal full matching schemes, which is one of the distance matching methods to re-weight the observations and has been implemented in the R *MatchIt* package. Briefly, optimal full matching assigned every treated and control unit to a single subclass, where each subclass consists of one treated unit and one or more control units, or vice versa. It is considered optimal when the subclasses and allocation of units are designed to minimize the total absolute distances (i.e., measured by propensity score) within subclasses in the matched sample. Weights are determined based on subclass membership, and the original propensity scores are used solely for the creation of subclasses and not directly to calculate the weights. The propensity score was created by logistic regression with program treated or not as dependent variable while age, gender, race, ethnicity, unweighted CCI, and one-year hospital utilization indices during the pre-intervention period (e.g., number of ED visits, number of inpatient visits, and number of outpatient visits) as independent variables.

We described the distribution of demographics, comorbidities, and pre-intervention health utilization in the intervention and control groups, respectively. For binary clinical outcomes (e.g., 30-day readmission post-intervention), we fit logistic regression models to estimate the intervention effect (i.e., exposure was program treated or not) and its standard error. The glm() function was used to fit the outcome with the matching weights (which were calculated from optimal full matching schemes) specified in the model. For continuous outcomes (e.g., LOS differences, difference in changes of 30-day readmission rate), we fit a linear regression model with the intervention or control as an independent variable along with the matching weights. Then, the avg_comparisons() function in the R *marginaleffects* package was used to estimate the cluster robust standard error of the effect size where subclasses have been generated during matching. All the association analyses with and without matching were conducted, and their results were compared. All the analysis was conducted by R 4.3.3, and p< 0.05 (two-sided) was considered a significant level.

## Results

In total, 100 patients were enrolled in the Healthy at Home study after 1,333 patients were invited (7.5% enrollment rate). After removing 12 patients with missing data on race or hospital utilization, 88 patients were analyzed in the intervention-treated group (mean age 67, 50% female). An additional 14,492 patients were included in the control group (Figure 1). Their characteristics are summarized in Table 1.

**Figure 1.**
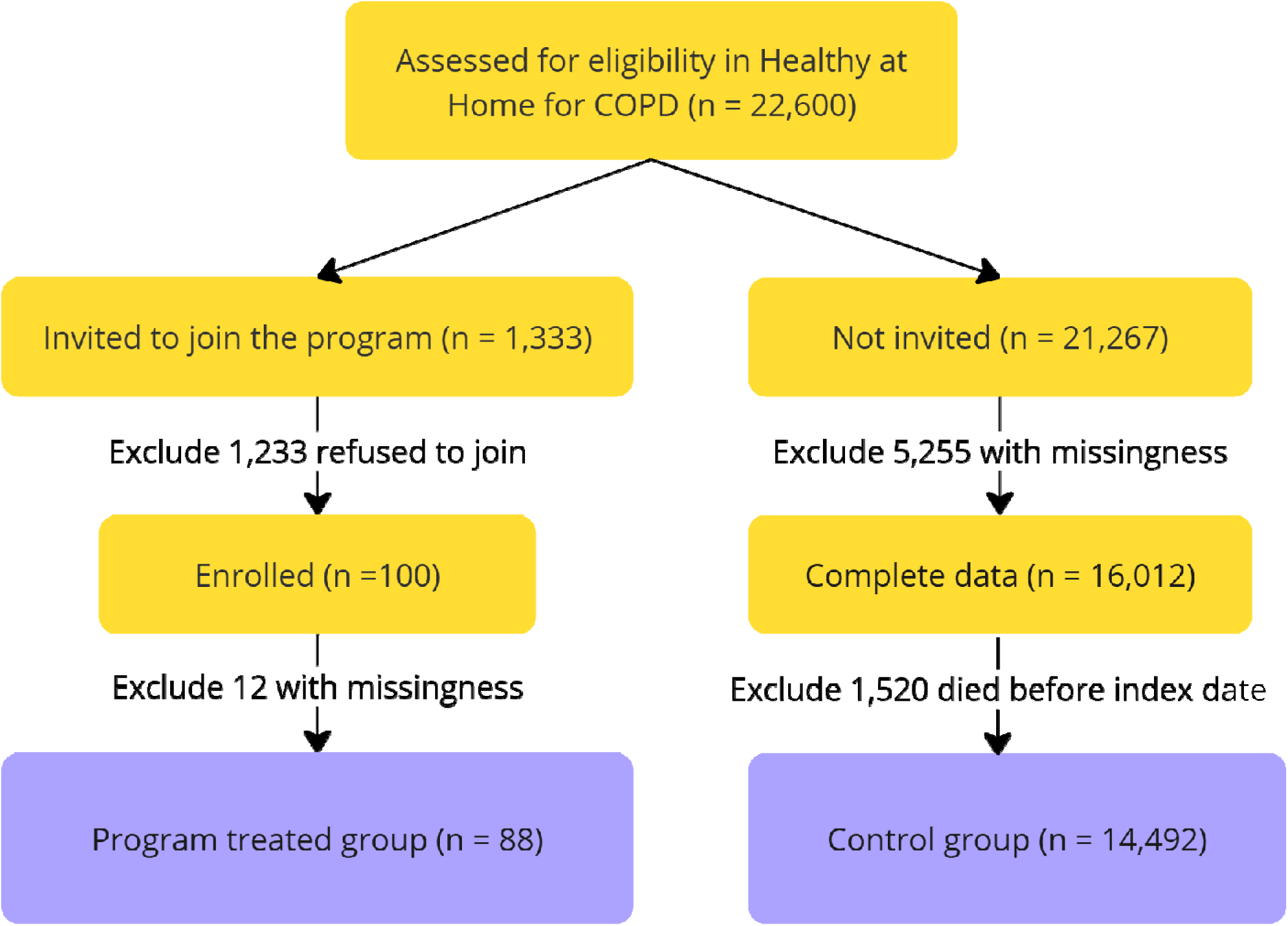
CONSORT Diagram of treated and control group.

**Table 1.** Characteristics of treated and control groups.

| Characteristics | Controls | Program treated |
| --- | --- | --- |
| n | 14492 | 88 |
| age (mean (SD)) | 69 (11) | 67 (10) |
| Male (%) | 6990 (48.2) | 44 (50.0) |
| race (%) |  |  |
| Black or African American | 301 (2.1) | 3 (3.4) |
| Other | 942 (6.5) | 7 (8.0) |
| White | 13249 (91.4) | 78 (88.6) |
| Hispanic or Latino (%) | 861 (5.9) | 6 (6.8) |
| Charlson Comorbidities index unweighted (mean (SD)) | 2.55 (1.79) | 2.56 (1.81) |
| Hospital utilization before index date |  |  |
| ED visits (mean (SD)) | 0.46 (1.53) | 0.32 (0.74) |
| Inpatient visits (mean (SD)) | 0.52 (1.15) | 0.51 (1.30) |
| Outpatient visits (mean (SD)) | 43.32 (54.79) | 81.20 (83.00) |
| Readmits 30 days (mean (SD)) | 0.34 (1.64) | 0.22 (0.67) |
| Readmits 90 days (mean (SD)) | 0.51 (1.96) | 0.40 (1.21) |
| Average LOS in days (mean (SD)) | 6.07 (12.18) | 8.01 (5.40) |
| Hospital utilization after index date |  |  |
| ED visits (mean (SD)) | 0.47 (1.67) | 0.32 (0.97) |
| Inpatient visits (mean (SD)) | 0.52 (1.15) | 0.51 (1.30) |
| Readmits 30 days (mean (SD)) | 0.40 (1.87) | 0.20 (0.98) |
| Readmits 90 days (mean (SD)) | 0.60 (2.20) | 0.38 (1.44) |
| Average LOS in days (mean (SD)) | 6.73 (12.90) | 5.81 (4.62) |
| Had 30 readmission after index date (%) | 2292 (15.8) | 6 (6.8) |
| Had 90 readmission after index date (%) | 2975 (20.5) | 12 (13.6) |
| Had ED visit after index date (%) | 3540 (24.4) | 18 (20.5) |
| Had inpatient visit after index date (%) | 4405 (30.4) | 23 (26.1) |
| Hospital utilization Changes (before minus after) |  |  |
| Change in ED visits (mean (SD)) | -0.01 (1.57) | 0.00 (1.05) |
| Change in inpatient visits (mean (SD)) | -0.09 (1.35) | 0.03 (1.10) |
| Change in readmits 30 days (mean (SD)) | -0.06 (1.72) | 0.01 (1.04) |
| Change in readmits 90 days (mean (SD)) | -0.09 (1.98) | 0.02 (1.37) |
| Change in average LOS (mean (SD)) | -0.66 (17.05) | 2.20 (7.07) |

The program-treated group of participants was comparably younger than the controls (mean age 67 vs. mean age 69). A higher overall percentage of non-White and Hispanic participants were recruited into the program. The mean number of outpatient visits before the enrollment date was also higher in the program-treated group than in the controls (81 visits vs. 43 visits). To better reduce the differences observed in the treated and control, the full optimal matching method upweighted younger, non-white, and more outpatient visits patients in the controls. The balance comparisons with and without matching are depicted in Table 2. Compared to the control group, the program-treated had a lower 30-day readmission rate (6.8% vs. 15.8%), lower 90-day readmission rate (13.6 % vs. 20.5%), and shorter LOS of inpatient visits (average LOS after index date: 5.8 vs. 6.7).

**Table 2.** Balance comparisons with and without matching.

|  | Program treated | Controls no matching | Controls weighted matching |
| --- | --- | --- | --- |
| n | 88 | 14492 | 14492 |
| Mean age | 67 | 69 | 66 |
| Male % | 50.0% | 48.2% | 51.3% |
| White % | 88.6% | 91.4% | 83.9% |
| Black or African American % | 3.4% | 2.1% | 5.2% |
| Other race % | 8.0% | 6.5% | 10.9% |
| Hispanic or Latino % | 6.8% | 5.9% | 10.5% |
| Mean unweighted CCI | 2.55 | 2.55 | 1.94 |
| Mean ED visits | 0.32 | 0.46 | 0.26 |
| Mean inpatient visit | 0.51 | 0.52 | 0.46 |
| Mean Outpatient visit | 81.20 | 43.32 | 62.38 |
| Total distance | 0.0102 | 0.006 | 0.0102 |

With the full optimal matching scheme, we detected a signal towards an overall decreased likelihood of any hospital admission (OR, 95% CI = 0.69 (0.44-1.03), p =0.07). We also found a significant association between intervention participation and reduced odds of 30 readmission: patients who joined the program were 61% less likely to have a hospital readmission within 30 days after enrollment (OR, 95% CI = 0.39 (0.16-0.95), p =0.04). Other clinical outcomes results are summarized in Table 3.

**Table 3.** Association of post-intervention clinical outcomes and program intervention.

| Outcome | Unadjusted |  |  | Optimal Full Matching |  |  |
| --- | --- | --- | --- | --- | --- | --- |
|  | OR | 95% CI | p | OR | 95% CI | p |
| Had 30 readmission | 0.39 | 0.17-0.89 | 0.026 | 0.39 | 0.16-0.95 | 0.038 |
| Had 90 readmission | 0.61 | 0.33-1.13 | 0.114 | 0.59 | 0.31-1.12 | 0.106 |
| Had ED visit | 0.80 | 0.47-1.34 | 0.388 | 0.70 | 0.43-1.15 | 0.158 |
| Had inpatient visit | 0.81 | 0.50-1.31 | 0.387 | 0.69 | 0.44-1.03 | 0.066 |
|  | Mean difference | 95% CI | P value | Mean difference | 95% CI | P value |
| LOS after index date | -0.92 | -8.92-7.09 | 0.821 | -1.23 | -5.4-2.94 | 0.564 |

When comparing the difference in differences between the treated and controls (Table 4), we observed a signal toward an overall reduction in health utilizations in the treated group. In particular, inpatient encounters decreased by 0.29 more than the controls (95% CI= −0.02-0.59, p= 0.07).

We also evaluated potential survival bias (i.e., unbalanced death rate between treated and controls) during the intervention: there were 6 patients died among the treated group (death rate = 6/88 = 0.07), while 1012 patients died in the controls (death rate = 1012/14492 = 0.07).

## Discussion

This study provides pilot data that a novel, community-based approach to COPD management integrating digital and mobile tools is directionally associated with reduced acute care utilization. While not statistically significant at the type-1 error rate of 5%, with the exception of 30-day readmission, we observed consistent directionality of intervention effects and quantified variance that can inform future effectiveness evaluations. Compared to a matched control group and their own pre-intervention data, participants experienced fewer hospitalizations, lower odds of 30-day readmission, and shorter mean inpatient length of stay. These findings build upon prior research demonstrating that this intervention enhances patient quality of life and reduces COPD-related distress. By leveraging unified digital and mobile technologies, this approach supports patients before and during acute exacerbations, addressing key accessibility challenges that often hinder timely COPD care. Further investigation is warranted to validate these pilot findings and assess the intervention’s potential as a scalable solution to the significant burden COPD places on patients, health systems, and communities. Our work aims to advance complementary digital-remote approaches to monitoring, evaluation, and treatment, creating a more comprehensive model for COPD care.

Previous studies have yielded mixed results on the effectiveness of digital health interventions intended to improve clinical outcomes for patients living with COPD.^27, 28^ There is similarly mixed evidence on mobile interventions that provide community-based in-person care to patients.^29^ Part of the limitations may be related to the siloing of resources: interventions that promote monitoring and self-management cannot respond expediently to clinical deterioration, and programs that provide mobile care have limited ability to know when to respond. No previous work has described a complementary program combining both digital and mobile components to provide not just monitoring and self-management support but also real-time proactive clinical response to address clinical deterioration when earlier signs and symptoms manifest in participants. Such an approach facilitates the execution of evidence-based strategies known to improve outcomes in COPD care-close monitoring, early symptom detection, and aggressive clinical intervention.^30^ Previous studies examining the use of highly structured disease-specific ambulatory clinics for COPD have shown that an aggressive, standard approach to COPD management decreases hospitalizations and ED visits; it stands to reason that a digital strategy that expands this approach to the community using digital and mobile resources would have a similar impact.^31^

The optimal full matching method we have used in Healthy at Home COPD is an emerging synthetic control method that can be used to evaluate the effectiveness of an intervention using external control data in many non-randomized clinical trials or single-arm trials.^32^ The optimal full matching falls at the intersection of matching, stratification, and weighting: it involves the formation of strata consisting of treated and control subjects; the analysis then incorporates weights that are derived from the stratification.^33^ We implemented the optimal full matching method here because of its two attractive features: one is that it is suitable for imbalanced sample sizes as we have a large number of controls relative to treated units. The second one is that the created matched subclasses are straightforward to interpret as it minimizes the total distance (e.g., propensity score difference) across all matched pairs or sets.

This pilot study has several limitations, including a small sample size and a nonrandomized design, necessitating the use of a weighted synthetic control group. As a result, the study may have been underpowered to detect smaller differences in acute care utilization and subgroup outcomes, particularly between participants who did and did not engage in telepulmonary rehabilitation. This limits the ability to determine whether the intervention’s effects were driven by telepulmonary rehabilitation alone or in combination with remote monitoring and mobile integrated health services. There may have been sampling bias that was not adequately adjusted for using a synthetic control approach. Although the study was designed to be decentralized, the recruitment strategy required significant participant activation, including reading digital messages, self-identifying eligibility, and opting into the study app. This likely influenced recruitment response rates. A more personalized recruitment approach, such as direct clinician referrals or in-person invitations, may have improved engagement. Future studies should consider refining recruitment strategies to enhance participation and ensure broader representation. Additional qualitative work is needed to further evaluate the acceptability of the Healthy at Home model amongst patients and clinicians. Finally, key implementation and sustainability measures, including cost-effectiveness, were not measured in this study. Describing the factors that will impact health systems and payors’ ability to integrate intensive community-based interventions into usual clinical practice and reimbursement models is key to ensuring the scalability of evidence-based interventions.

Further research should evaluate the use of a hybrid digital-mobile intervention for COPD exacerbation like Healthy at Home in a fully powered randomized controlled trial in order to evaluate its effectiveness in real-world practice. In addition to effectiveness measures, implementation measures, including cost-effectiveness, should be rigorously measured. Additionally, additional qualitative evaluation with key stakeholders, including patients, clinicians, and health system leaders to evaluate the intervention’s acceptability, sustainability, and translatability from research study into clinical practice. Finally, adaptations to the intervention itself to improve its effectiveness, such as leveraging a more proactive approach to clinical evaluation when biometric changes are detected, should be considered. Healthy at Home ideally leverages the flexibility, mobility, and accessibility of digital tools and mobile care delivery assets to provide comprehensive care to patients in the community, effectively eliminating many barriers to treatment by literally bringing healthcare to where patients are. It expands the value of siloed tools to enhance the utility of modern health technology and assets to facilitate high-quality treatment and mitigate longstanding disparities in care.

## Data Availability

All data produced in the present study are available upon reasonable request to the authors

