## Supplemental Table 1 for "Evaluation of an Integrated Digital and Mobile Intervention for COPD Exacerbation"

| **Table 1: Healthy at Home Intervention Components** | | | |
| --- | --- | --- | --- |
| **Intervention Component** | **Description** | **Interaction with other components** | **All Participants or Opt-In** |
| MyDataHelps App (main study smartphone app) | Houses all participant-facing forms and questionnaires including screening and consent. Prompts participants to complete scheduled assessments. Receives information from biosensors and is linked to participants’ EHR and claims data to aggregate all data streams. Generates momentary assessments triggered by participant responses or biometric data. | All participant-level MyDataHelps data is visible to the tele-pulmonary-rehabilitation and MIH team on the study dashboard. Clinical data can trigger alert to participants through the app suggesting a home assessment from the MIH team and connect participants directly with the MIH visit request-line. | All |
| Fitbit Smartwatch (and Fitbit app) | Collects biometric data. All data is visible to participants, study team, MIH clinicians and tele-pulmonary-rehabilitation coaches through the MyDataHelps. | Biometric patterns trigger alert to patients suggesting MIH visits. Biometric data is visible to clinical and research teams. | All |
| Mobile Integrated Health Program (community paramedics and MIH physicians) | Community paramedics are available on-demand to perform home visits to evaluate and treat participants experiencing acute symptoms with support from a supervising physician. The program is specifically equipped to initiate treatment for COPD exacerbation. | MIH team can view participant level data in MyDataHelps. MIH visits offered to participants reporting worsening symptoms or exhibiting concerning biometric data through the study app. Research and tele-pulmonary-rehabilitation team refer participants to MIH team for all acute clinical concerns. | All |
| Wellinks Virtual Pulmonary Rehabilitation Program | Coaching program to support participant education, treatment-adherence, and goal setting, as well as a virtual pulmonary rehabilitation. Hosted on a separate program-specific app. The Wellinks staff is comprised of registered nurses, respiratory therapists, and certified health and wellness coaches. | Tele-pulmonary-rehabilitation team can view participant level data in MyDataHelps to support coaching plans. tele-pulmonary-rehabilitation team can contact MIH team with any acute concerns requiring clinical evaluation. | Opt-In |
| Wellinks equipment kit (spirometer, pulse oximeter, exercise band) | Used with the tele-pulmonary-rehabilitation team to support pulmonary coaching plan and collect additional relevant biometric data. | Spirometry and pulse oximetry data is integrated with the MyDataHelps app and is visible to the study and clinical teams. | Opt-In |
